# “Men are the main COVID-19 transmitters: lessons from couples”

**DOI:** 10.1101/2021.08.18.21262187

**Authors:** Monize V. R. Silva, Mateus V. de Castro, Maria Rita Passos-Bueno, Paulo A. Otto, Michel S. Naslavsky, Mayana Zatz

## Abstract

**Background:** COVID-19 has affected millions of people worldwide. Clinical manifestations range from severe cases with a lethal outcome to mild or asymptomatic cases. Although there is the same proportion of infected genders, men are more susceptible to severe COVID-19, with a higher risk of death than women. This sex-bias may be explained by biological pathways.

**Methods:** We performed an epidemiological survey from July 2020 to July 2021 including 1744 unvaccinated adult Brazilian couples with at least one infected spouse despite living together during the COVID-19 infection without protective measures. The presence or absence of infection was confirmed by RT-PCR and/or serology results. The couples were divided between groups where both partners were infected (concordant couples) or only one spouse remained asymptomatic despite the close contact with the infected one (discordant couples). Statistical analysis of the collected data was performed aiming to verify a differential transmitter potential between genders in household contact.

**Results:** The combination of our data collected from concordant and discordant couples showed that the man is the first (or the only) affected in the major occurrences when compared to women. Our findings support other published surveys and are in concordance with previous studies of our group.

**Conclusions:** These observations support the hypothesis according to which male individuals are more efficient virus transmitters than females, independently of the use of protective masks. In short, the present study confirmed the existence of gender differences not only for susceptibility to infection and resistance to COVID-19 but also in the transmission rate.

**HIGHLIGHTS:**

- There are sex differences in COVID-19 susceptibility and transmission between couples with direct contact without protective measures;
- Men are more efficient virus transmitters than women;
- Sex-bias in COVID-19 transmission can be explained by differences in viral load in saliva, immune response and also behavioural protective differences between genders.

## INTRODUCTION

Since the first reported case of COVID-19 caused by SARS-CoV-2 infection in Wuhan, China, in late December 2019, about 200 million of individuals globally have been infected by the novel coronavirus reaching the mark of 4 million recorded deaths. Under this tragic scenario, efforts have been conducted to better understand the disease and its impacts in public health and in various aspects of human life. Thus, investigating the factors involved in the transmission rates to ultimately slow down the spread of the virus, are of utmost relevance. In this sense, many scientific studies have examined the transmission dynamics of COVID-19, by tracing the route of transmission through human-to-human contact, monitoring or evaluating the role that some environmental factors may play in facilitating the spreading rate of the disease (1).

Although there are no gender differences in the proportion of people infected with SARS-CoV-2, men are more susceptible to severe COVID-19 and death (2, 3). Independently of age, they are more likely to have complications by COVID-19 than women and if hospitalization is required, males have more risk of death than females (4). Interestingly, comparable gender differences also occur for other viral infections (5).Furthermore some behavioural aspects such as the adoption of COVID-19 prevention and control measures vary between genders. A survey conducted in March-April 2020 indicated that men are more reluctant than women to wear protective masks and respect social distancing (6).

It has been suggested that some individuals named “superspreaders” could transmit the virus to a great number of persons. However, it is not known if this could be explained by behaviour (men speaking louder without mask) or biologically (differences in lung capacities between sexes and ages). Thus, viral transmission capability could be influenced by less aerosol emission by female and children (7).

Sex-based differences may be also associated with variances in biological pathways, such as immune responses against SARS-CoV-2 and/or expression of X-chromosome–encoded genes (7). These observations led us to question whether the virus could be transmitted more frequently by men than women, independently of protection measures. In order to circumvent behavioural differences we analysed the virus transmission in couples who did not keep conjugal distancing during the infection period neither the use of protective measures.

In our previous genetic study of 81 discordant couples for COVID-19, where one person was infected and symptomatic and the partner remained asymptomatic and serum-negative (despite remaining in close contact and sharing the same bed throughout the disease), we observed that there were significantly more women in the asymptomatic group (8). Intriguingly, we noticed several reports of couples (particularly where both are physicians) where the wife was infected by SARS-Cov-2 and clinically affected while the husband remained asymptomatic and some months later, the previously asymptomatic husbands got infected and symptomatic after contact with a male patient. These observations led us to investigate the household transmission dynamic between couples through a questionnaire to identify the first infected spouse among couples.

## METHODS

Here we present an epidemiological survey with adult Brazilian couples (mean age of 45 years old) who were living together during the COVID-19 infection event without protective measures. From July 2020 to July 2021, we collected data from e-mails and questionnaires of 2048 Brazilian couples where at least one spouse was infected by SARS-CoV-2 and symptomatic. The positive diagnosis in symptomatic individuals was confirmed by RT-PCR and/or serology for the infected partners while negative results in both tests were confirmed in non-infected partners. From this total number, we selected non-redundant 1744 couples that presented completed information related to the infection event, sex, age, and diagnostic tests results. Chi-squared tests were applied to couples from e-mails and questionnaires, showing that their corresponding data were homogeneous and thus could be assembled in two larger groups : a) concordant couples where one spouse who had confirmed COVID-19 transmitted the infection to the partner (chi-squared test value of χ^2^ = 0.0309, 1 d.f., P = 0.8605) and b) discordant couples where one spouse was infected by SARS-CoV-2 while the partner was not, as confirmed by negative RT-PCR and serology results after viral exposure(chi-squared test value of χ^2^ = 0.5162, 1 d.f., P = 0.4724).Then, the concordant and discordant couples were subdivided according to which partner was infected first: men to women; men to men; women to women and women to men. Our whole data collection rationale from partners is presented in **Figure 1**. The calculated chi-squared test values (all with 1 d.f.) obtained to verify the null hypothesis of equality of gender frequencies between concordant and discordant couples are listed on **Table 1**, together with the corresponding numbers of subjects.

**Figure 1.**
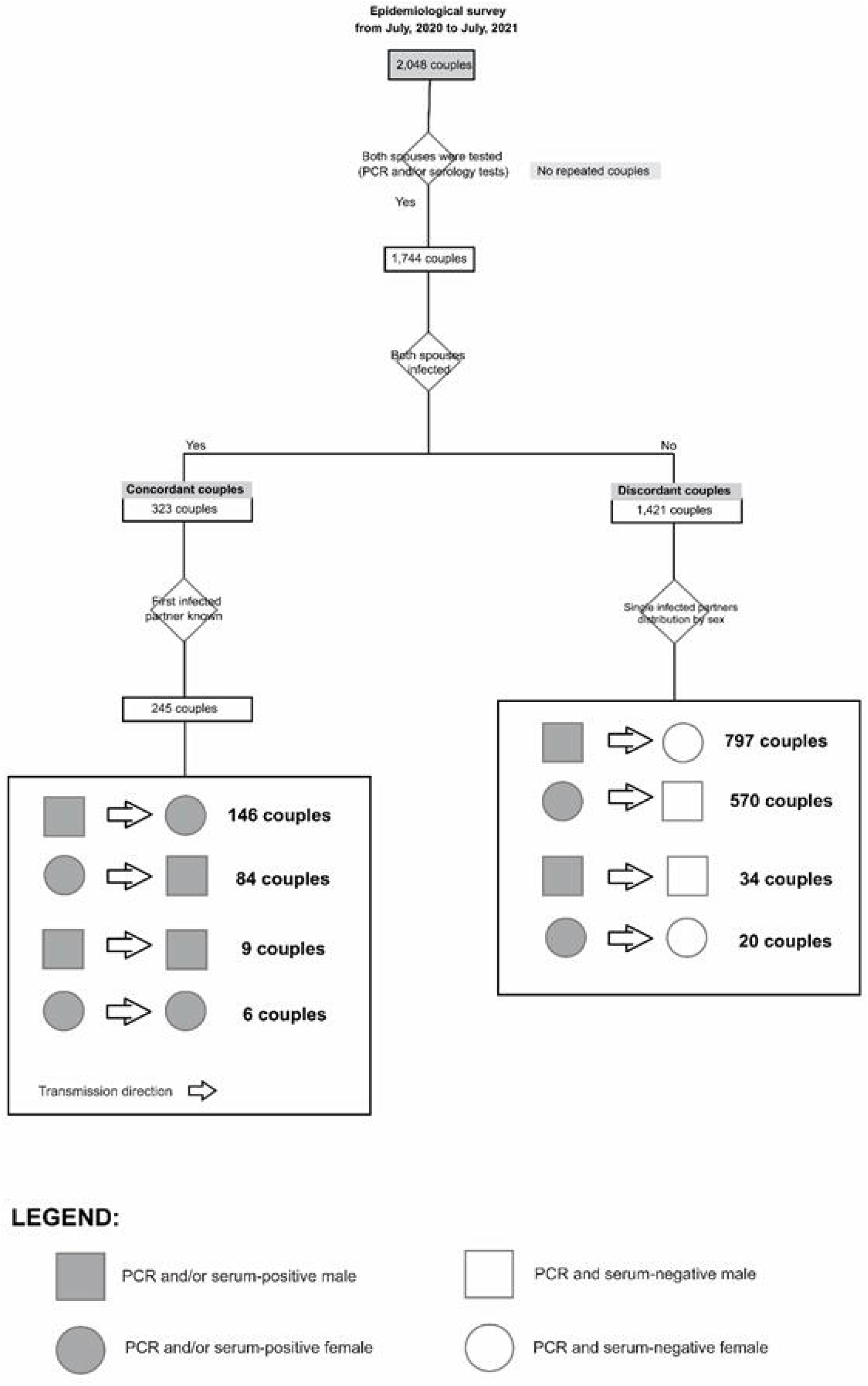
Survey data collection rational diagram.

**Table 1.** Numerical description of couple groups composition and chi-square (χ^2^) values.

| Group | Men | Women | Total | Chi-square ($\chi^2$) values |
| --- | --- | --- | --- | --- |
| Discordant | 797 | 570 | 1367 | 37.6950 |
| Concordant | 146 | 84 | 230 | 16.7130 |

## RESULTS AND DISCUSSION

Analysis of the group of concordant couples as to COVID-19 serological symptomatology showed that men are significantly more infectious than women, with an estimated chance of 146/230 = 0.635 (exact 95% confidence interval 0.569 - 0.697) of being the first affected among concordant couples. The corresponding female values were 84/230 = 0.365 with an exact 95% confidence interval of (0.303 - 0.431). These differences in male and female values are statistically significant (χ^2^ = 16.713, 1 d.f., P <<0.0005 for the concordant group.

The group of the COVID-19 discordant partners showed again that men are preferentially affected, with a probability value estimated as 797/1367 = 0.583 with a 95% confidence interval of (0.557 - 0.609). The corresponding female values were 570/1367 = 0.417 and (0.391 - 0.443). The hypothesis of an equal distribution of males and females is excluded by this setup (χ^2^ = 37.695, 1 g.l., P<<0.0005).

Since the number of homosexual couples was relatively small, they were not directly considered in our statistical tests to estimate the gender distribution of the infection and resistance capabilities. However, we observed that they are in perfect accordance with those obtained from the rest of our sample, since among the homosexual couples from our survey where both were infected by SARS-CoV-2, nine are male and six female. Among 37 discordant homosexual couples from our sample, the affected is male and in 25 the affected member is a female. The chi-square test used for verifying the null hypothesis of equality of sexes had a value o fχ^2^ = 3.522 (1 d.f.), P = 0.061, suggesting again a tendency for a significant excess of males. Although these data do not allow analysing gender transmission, both observations indicate a larger degree of susceptibility of male individuals to COVID-19 infection.

Using a global approach of combining the data from concordant and discordant heterosexual couples, we could determine the sex-specific capabilities of infection and resistance. Indeed, we verified that the man is the first (or the only) affected in 146 + 797 = 943 occurrences and the woman in 84 + 576 = 660 instances (χ2 = 49.962, 1 g.l., P <<0.0005).

All the results obtained in the present study strongly suggest that male individuals not only are more susceptible to COVID-19 severity, as shown in worldwide literature, but they also more likely to transmit the virus to their partners when compared to females in the household transmission context. The epidemiological findings in the present survey are consistent with the results of other published surveys involving couples where one of the partners was infected by their spouses (10, 11). Female individuals aged between 17 and 65 years are also frequently indicated to be secondary cases (11). Similar evidence of male individuals as the main source of transmission was previously reported for *Mycobacterium tuberculosis* Infection (12).

One of the possible current hypotheses for such gender variable transmission rate is a differential viral load in saliva, which has been explored as an important clinical measure of disease severity due to its positive association with many COVID-19 inflammatory markers (13).

Interestingly, a recent study of our group (14) observed that, although there were no observed gender differences in viral load in nasopharyngeal samples, adult males showed a significantly higher viral load in saliva samples (verified by RT-LAMP viral testing) than females of comparable ages.

These observations, together with the evidence of higher aerosol emission by men which make them more likely to be “super-spreaders” than women, support the hypothesis according to which male individuals are more efficient virus transmitters than females, independently of the use of protective masks.

## CONCLUSION

In short, the present study confirmed the existence of gender differences not only for susceptibility to infection and resistance to Covid-19 but also in the transmission rate.

## Data Availability

Individual level data can be shared by authors upon reasonable request.

## ACKNOWLEDGMENTS AND FUNDING

The authors are extremely grateful to all volunteers for their participation and collaboration. This work was supported by (FAPESP/Brazil) [grant numbers 2013/08028-1, 2014/50931-3 and 2020/09702-1], the National Council for Scientific and Technological Development (CNPq) [grant number 465355/2014-5] and JBS S.A [grant number 69004].

## ETHICS DECLARATION

This study was approved by the Committee for Ethics in Research of the Institute of Biosciences at the University of São Paulo (CAAE 34786620.2.0000.5464).

## Notes

### Competing Interest Statement

The authors have declared no competing interest.

### Clinical Trial

This is a retrospective epidemiological survey with adult Brazilian couples (mean age of 45 years old) who were living together during the COVID-19 infection event without protective measures.

### Author Declarations

This study was approved by the Committee for Ethics in Research of the Institute of Biosciences at the University of Sao Paulo (CAAE 34786620.2.0000.5464).

## REFERENCES

1. Correa-Araneda F, Ulloa-Yáñez A, Núñez D, Boyero L, Tonin AM, Cornejo A, et al. Environmental determinants of COVID-19 transmission across a wide climatic gradient in Chile. Scientific Reports. 2021;11(1).

2. Peckham H, de Gruijter NM, Raine C, Radziszewska A, Ciurtin C, Wedderburn LR, et al. Male sex identified by global COVID-19 meta-analysis as a risk factor for death and ITU admission. Nature Communications. 2020;11(1).

3. Dehingia N, Raj A. Sex differences in COVID-19 case fatality: do we know enough? The Lancet Global Health. 2021;9(1):e14–e5.

4. Lazzeri C, Vahidy FS, Pan AP, Ahnstedt H, Munshi Y, Choi HA, et al. Sex differences in susceptibility, severity, and outcomes of coronavirus disease 2019: Cross-sectional analysis from a diverse US metropolitan area. Plos One. 2021;16(1):e0245556.

5. Klein SL, Flanagan KL. Sex differences in immune responses. Nature Reviews Immunology. 2016;16(10):626–38.

6. Galasso V, Pons V, Profeta P, Becher M, Brouard S, Foucault M. Gender differences in COVID-19 attitudes and behavior: Panel evidence from eight countries. Proceedings of the National Academy of Sciences. 2020;117(44):27285–91.

7. Lewis D. Superspreading drives the COVID pandemic — and could help to tame it. Nature. 2021; 590(7847):544–6.

8. van Luzen JA, Marcus. Sex differences in infectious diseases - common but neglected. Journal of Infectious Diseases. 2014 2014;209.

9. Castelli EC, de Castro MV, Naslavsky MS, Scliar MO, Silva NSB, Andrade HS, et al. 2021.

10. Qifang Bi JL, Isabella Eckerle, Stephen A. Lauer, Laurent Kaiser, Nicolas Vuilleumier, Derek A. T. Cummings, Antoine Flahault, DusanPetrovic, IdrisGuessous, Silvia Stringhini, Andrew S. Azman&SEROCoV-POP Study Group. Insights into household transmission of SARS-CoV-2 from a population-based serological survey. Nat Commun. 2021;12.

11. Hall Jah, R. J.; Zaidi, A.; Woodhall, S. C.; Dabrera, G & Dunbar, J. K. HOSTED - England’s Household Transmission Evaluation Dataset: preliminary findings from a novel passive surveillance system of COVID-19. International Journal of Epidemiology. 2021 09 April 2021;50(3):743-52. Epub 752.

12. Silva J, Lucas C, Sundaram M, Israelow B, Wong P, Klein J, et al. Saliva viral load is a dynamic unifying correlate of COVID-19 severity and mortality. Preprint. 2021.

13. Kobayashi Gsb, L. A.; Moreira, D. P.; Suzuki, A. M.; Hsia, G. S. P.; Pimentel, L. F.; Paiva, A. P. B.; Dias, C. R.; Lourenco, N. C. V.; Oliveira, B. A.; Manuli, E. R.; Corral M. A.; Cavacana, N.; Mitne Neto, M.; Sales, M. M.; Dell’ Aquila, L. P.; Razuk Filho, A.; Parrillo, E. F.; Mendes-Correa, M. C.; Sabino, E. C.; Costa, S. F.; Leal, F. E.;Sgro, G. G.; Farah, C. S.; Zatz, M.; Passos-Bueno, M.. A novel RT-LAMP workflow for rapid salivary diagnostics of COVID-19 and effects of age, gender and time from symptom onset. Diagnostics. 2021 (in press)

